# Physical Therapy Utilization and 12-Month Pain and Functional Improvement in Patients Undergoing Arthroscopic Rotator Cuff Repair Surgery: A Case Series

**DOI:** 10.64898/2026.02.19.26346640

**Authors:** Chris J. Pierson, Brady Moore, Tristan Elias, Joshua C. Harris, Jeremy Somerson

**Author notes:** Corresponding Author Chris J. Pierson, Mailing: Research Building 17, 301 University Blvd, Galveston, TX, 77550.

## Abstract

**Background:** Arthroscopic rotator cuff repair (RCR) is a common surgical intervention used to address rotator cuff-related pain after other conservative interventions have been exhausted. With a continuing increase in procedures, evidence-based outcomes research is needed to identify key parameters of postoperative rehabilitation planning.

**Objective:** We aim to identify rehabilitation planning factors leading to better outcomes while also providing clinicians with reference data to describe the magnitude of pain and functional improvement following RCR.

**Methods:** For this observational study of patients undergoing RCR surgery and physical therapy, demographic variables and patient-reported outcome measures (PRO) were collected preoperatively and up to 12 months postoperatively Four multiple linear regression models were created, one for 12-month Visual Analog Scale (VAS) score, the second for VAS improvement, the third for 12-month American Shoulder and Elbow Surgeons (ASES) function score, and the fourth for ASES function improvement.

**Results:** The 29 participants had a median age of 62 years, median baseline VAS of 4.9, ASES composite score of 45, and Veterans-Rand 12 Mental Component Score of 53.5. In univariate analysis, one variable was associated with 12-month VAS score and two were associated with 12-month VAS improvement. No associations were found with 12-month ASES function score, and one variable was associated with ASES function improvement. With our sample, multivariable analyses provided no significant association or predictor for VAS or ASES function scores.

**Conclusions:** Our hypothesis was not supported, and we did not find an association between physical therapy wait time prior to evaluation or visit frequency and PRO measures or improvements. We observed that 12-month PRO measures and improvements can be predicted using baseline measures among this population.

## INTRODUCTION

Arthroscopic rotator cuff repair (RCR) is a common surgical intervention used to address rotator cuff-related pain after other conservative interventions have been exhausted. There were 151,866 arthroscopic RCR procedures covered by private insurance in the United States from 2004 to 2009, with approximately 14 procedures for every 1,000 patients with an orthopaedic International Classification of Diseases, Ninth Revision (ICD-9) code.^1^ Given the increase in surgical procedures, there is an expanding need for evidence-based rehabilitation strategies to optimize patient outcomes.^2^ A recent population-based study of commercially insured patients following RCR found that 81% utilized formal rehabilitation; of that group, 94% worked with a physical therapist for postoperative rehabilitation.^3^

Although the benefit of postoperative rehabilitation is well-established,^4–6^ the optimal rehabilitation framework and timing of its initiation are topics of debate.^7–9^ The dosage and timing written in most rehabilitation protocols are based on clinician experience and expert opinion rather than evidence-based rationale, resulting in substantial variation among providers.^4^ The goal of postoperative rehabilitation after RCR is to promote tendon healing and protect the repair while simultaneously restoring function and range of motion.^10^ Several studies have been published describing the factors leading to utilization of therapy, but these did not investigate the associations between utilization and treatment outcomes.^3,11,12^ Prior research has found physical therapy (PT) utilization rates following RCR to be 80% to 82% in a commercially insured population.^3,10^ These studies have examined PT attendance as an outcome of interest, but not as a predictor of treatment outcomes.

The first objective of this study was to identify the rehabilitation treatment plan factors within 6 months of surgery that are associated with 12-month scores as well as 12-month improvements in pain and American Shoulder and Elbow Surgeons (ASES) functional score. Identifying the associated utilization factors can help patients, physicians, and physical therapists allocate resources to create a multidisciplinary plan of care that will return the best value. We hypothesized that after accounting for significant covariates, patients with greater outcome improvements had a shorter PT evaluation wait time and attended more PT visits.

The second objective was to provide clinicians with reference data for patient education regarding the magnitude of pain and functional improvement following RCR. Providing patient-accessible data to guide surgical outcome expectations by timepoint may help with patient satisfaction and improve patient-provider trust.

## METHODS

This study received Institutional Review Board approval (IRB# 20-0306) and followed the STROBE guidelines for observational studies (Appendix A).^13^ In this retrospective cohort analysis of a prospectively maintained patient-reported outcome database, surgical procedures performed by a single fellowship-trained shoulder and elbow surgeon between April 6, 2018 and October 13, 2021 were analyzed. These clinics are at a large, urban, tertiary medical center in the Southern United States. Patients were included if they underwent arthroscopic rotator cuff repair surgery (Current Procedural Terminology [CPT] code 29827), were treated at our institution’s affiliated PT clinics, and had data available for both outcome variables and all 10 assessed variables. Patients with revision surgeries and patients in the criminal justice system were excluded. Intraoperative description of tear size was recorded by the surgeon in the operative note.

All patients underwent a routine preoperative interscalene block followed by general endotracheal anesthesia. Arthroscopic RCR was performed in the beach chair position in all cases. Patients were routinely discharged on the day of surgery. At discharge, patients received a standardized prescription of ibuprofen 800 mg 3 times daily (unless contraindicated), ondansetron as needed for nausea, docusate for stool softener, tramadol 50 mg, and hydrocodone/acetaminophen 5/325 mg for breakthrough pain.

A standardized PT protocol was provided to all physical therapists by the treating orthopedic surgeon. Patients with small tears (≤ 3 cm anteroposterior width) without retraction were treated with single-row repairs followed by sling immobilization and passive range of motion for 4 weeks. Active range of motion and light resistance exercises were initiated at 4 weeks, and progressive strengthening added at 10 to 12 weeks. Patients with larger and/or retracted tears were treated with double-row repairs and had sling immobilization maintained for 6 weeks with supported passive table slides started at 2 weeks. Active range of motion was initiated at 6 weeks and strengthened at 12 weeks.

Demographic variables, clinical variables, and PT utilization data were extracted from electronic medical record chart review. The days from surgery to initiation of physical therapy evaluation were recorded as in prior research.^3^ The number of visits within 6 months of surgery were counted and categorized based on multiples of 6 to reflect average monthly frequency. This was categorized as (1) 1 to 5 visits, (2) 6 to 11 visits, (3) 12 to 17 visits, (4) 18 to 23 visits, (5) 24 to 30 visits, or (6) more than 30 visits. These categories represent 1, 2, 3, 4, or more than 5 visits per month, respectively. The 6-month utilization range was chosen because this is within Phase 4 of recovery, where strengthening is typically the goal,^14^ and many patients can continue this phase independently with their home exercise program.

### Outcome Variables

The ASES score is a PRO recommended by the American Academy of Orthopaedic Surgeons (AAOS) for use with rotator cuff injuries.^15^ This facilitates team-based treatment by addressing domains of pain management and specific functional tasks.^16^ The composite score of 100 consists of a pain score from 0 to 50 and a functional activity score from 0 to 50. This functional activity score is derived from a set of responses summed from a 30-point response and converted to a score from 0 to 50. This has shown strong psychometric properties for patients with shoulder dysfunction, with acceptable test-retest reliability and internal consistency values.^17^ A higher score is better, the minimal detectable change is 9.7 ASES points, and the minimal clinically important difference is 6.4 ASES points.^17^ This was provided on paper or via emailed survey link to study participants prior to and at pre-specified timepoints following surgery.

Visual analog scale (VAS) scores, which provide another patient-reported metric specifically for patient pain experiences and subsequent change following treatment, are also recommended by the AAOS.^15^ VAS is scored on a 0 to 10 scale where 0 indicates no pain and 10 indicates the patient’s worst pain imaginable. Patients reported this on a paper analog scale or via an emailed survey link with a slider function.

### Other Model Factors

Several non-modifiable demographic and clinical factors are known to affect PRO measures after RCR. Although success can be defined in different ways, multiple publications support the idea that younger patients have better outcomes.^18–22^ Conflicting evidence exists for sex, as female sex is associated with greater frequency of high levels of pain 6 weeks following surgery.^23^ Another study found 7- and 90-day VAS pain levels to be the same among sexes.^24^ Patients with a Veterans RAND 12-item Health Survey Mental Component Summary score (VR-12 MCS) higher than 40.00 have been shown to have significantly faster improvements in pain and function.^25^ Having a preoperative ASES score over 63 is associated with less relative improvement, but higher final ASES scores.^26^ Preoperative pain at rest is associated with greater frequency of pain 6 weeks following surgery.^23^ A larger tear size, identified intraoperatively, has a negative effect on tendon healing^19,21^ and more retears,^27^ but interestingly, smaller tears are associated with more frequent and severe postoperative pain.^27^ At our institution, the PT protocol provided is different between small and large tear sizes.

### Statistical Analysis

Statistical analyses were performed using SAS 9.4 (SAS Institute Inc., Cary, NC). Univariate analyses were performed for each independent variable to 12-months and improvement in VAS and ASES function as dependent variables. In univariate analyses, Kruskal-Wallis tests were used for categorical variables and simple linear regressions for continuous variables. The Shapiro-Wilk test was used to assess distribution for each dependent variable. VR-12 MCS scores and number of PT visits were assessed twice each, once categorical and once continuous.

Four combined multivariable linear regression models were created with all independent variables to assess which were associated with 12-month VAS score, 12-month VAS improvement, 12-month ASES functional score, and 12-month ASES functional improvement. Levels of statistical significance were set at *P* = 0.05, with levels of clinical significance at ASES minimal clinically important difference (MCID) of 6.4 points^17^ and VAS MCID of 2.5 points.^28^

Prior to analysis, the 12-month VAS improvement (Shapiro-Wilk W = 0.97, *P* = 0.5727) and 12-month ASES functional improvement (W = 0.96, *P* = 0.4830) both demonstrated a normal distribution. Additionally, both 12-month dependent variables had a significantly skewed distribution, with VAS strongly clustered around the minimum of 0 (W = 0.74) and ASES function (W = 0.77) strongly clustered around the maximum 30. Due to the non-normal distribution, we performed comparisons using non-parametric tests.

## RESULTS

Sixty consecutive patients receiving surgery from April 6, 2018 to October 13, 2021 were assessed for inclusion. Of these, 18 did not have 1-year of follow-up data for VAS, 23 did not have ASES scores, 11 did not attend at least 1 PT visit at our institution, 1 had a revision surgery, and 1 was deceased. Complete VAS data for analysis were available for 29 patients, and complete ASES function data were available for 24 patients. Descriptive statistics of subject characteristics, PT utilization data, and clinical outcomes can be found in Table 1 and Table 2.

**Table 1.** Subject Categorical Data (n = 29)

|  | N (%) |
| --- | --- |
| Male sex | 19 (66) |
| Race |  |
| Asian | 0 |
| Black or African American | 3 (10) |
| More than 1 race | 0 |
| Unknown | 0 |
| White | 26 (90) |
| Ethnicity |  |
| Hispanic or Latino | 3 (10) |
| Non-Hispanic or Latino | 26 (90) |
| Baseline Veterans-RAND 12<br>Mental Component Score |  |
| Above 40 | 23 (79) |
| Below 40 | 6 (21) |
| Tear size |  |
| Small | 17 (59) |
| Large | 12 (41) |
| Physical Therapy visit frequency |  |
| One visit per month | 4 (14) |
| Two visits per month | 8 (28) |
| Three visits per month | 4 (14) |
| Four visits per month | 8 (28) |
| Five visits per month | 3 (10) |
| Six visits per month | 2 (7) |

**Table 2.** Subject Continuous Data (n = 29)

|  | **Mean ± SD** | **Median (IQR)** | **Min-Max** |
| --- | --- | --- | --- |
| Age, years | 59.2±9.3 | 62 (10) | 38-72 |
| Baseline VAS pain level | 4.9±2.1 | 4.9 (3.6) | 0.76-8.7 |
| Baseline ASES function score | 12.1±4.8 | 11 (3.0) | 4.0-21.0 |
| Baseline ASES composite score | 44.69±12.9 | 45 (16.57) | 13.6-71.32 |
| Baseline VR-12 MCS | 50.4±12.1 | 53.4 (15.5) | 26.6-67.9 |
| Physical therapy evaluation wait time, days | 20.8±10.4 | 19 (13) | 2.0-48.0 |
| Number of physical therapy visits | 15.4±8.7 | 15 (13) | 3.0-31.0 |
| 12-month VAS pain level (n=29) | 1.5±2.1 | 0.4 (1.4) | 0-7.8 |
| 12-month VAS improvement (n = 29) | 3.5±2.3 | 2.9 (3.2) | 0.5-8.4 |
| 12-month ASES Function (n=24) | 24.9±6.5 | 28 (9) | 11-30 |
| 12-month ASES Function improvement (n = 24) | 12.0±7.6 | 12.0 (14.0) | -3.0-25 |

ASES = American Shoulder and Elbow Surgeons, IQR = interquartile range, VAS = visual analog scale, VR-12 MCS = Veterans RAND 12-Item Health Survey Mental Component Score

In univariate analysis, a simple linear regression found the 12-month VAS improvement to be highly associated with the 12-month ASES improvement (F = 139.21, *P* < 0.0001, R^2^ = 0.84). Interestingly, 12-month ASES function scores were not associated with 12-month VAS scores (F = 0.11, *P* > 0.7398).

### 12-Month VAS Score and ASES Function Score

Detailed univariate analyses results can be found in Table 3. In univariate analysis, the 12-month VAS score was significantly associated with only baseline VAS. A higher baseline VAS, indicating worse pain, is associated with a higher 12-month VAS score, indicating still worse pain (F = 5.35, *P* = 0.0286). At the individual level, a higher 12-month VAS score of 0.4 can be predicted by every 1.0 point increase in baseline ASES scores. With univariate analyses, there were no significant predictors of 12-month ASES function scores.

**Table 3.**
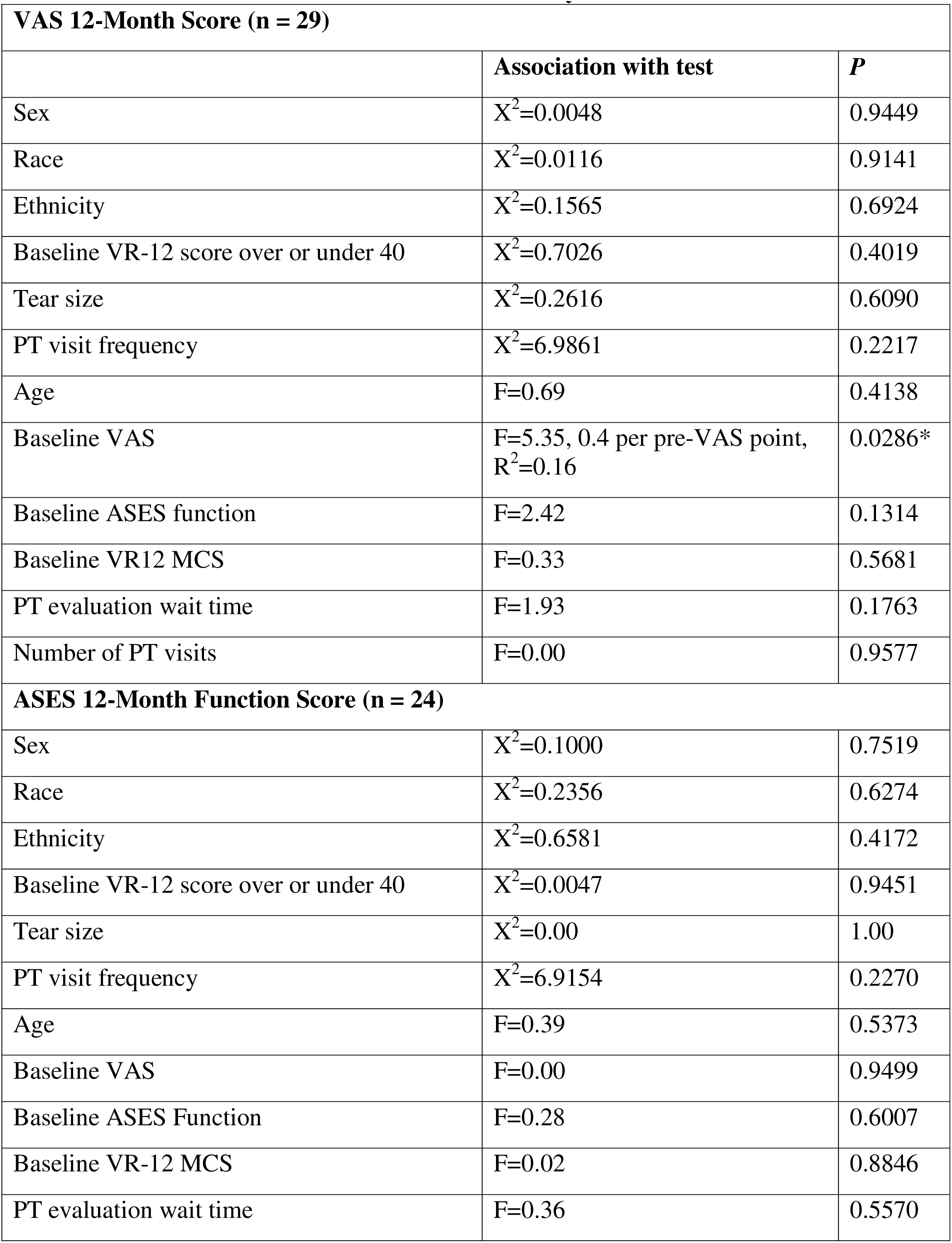
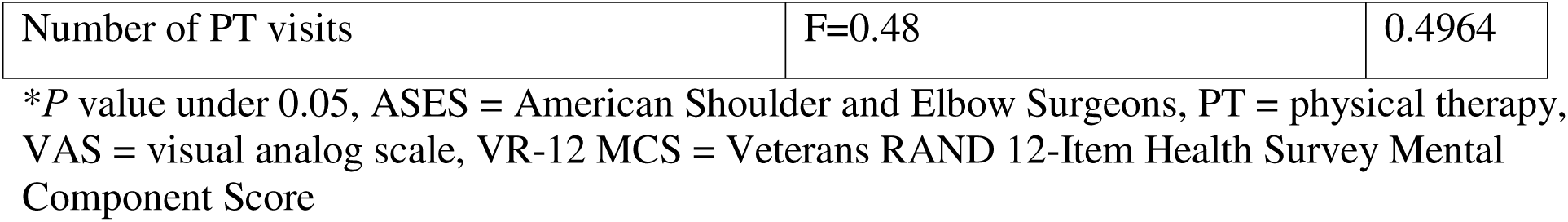
Results of 12-Month Score Univariate Analyses.

### 12-Month Improvements in VAS Score and ASES Function Score

Detailed univariate analyses results can be found in Table 4. In univariate analysis, the 12-month VAS improvement was significantly associated with baseline VAS (F = 12.19, *P* = 0.0017). With higher baseline pain, more pain improvement is predicted. At the individual level, a greater 12-month VAS improvement of 0.6 can be predicted by every 1.0 point increase in baseline VAS.

**Table 4.** Results of Univariate Analyses for 12-Month Improvements.

| **VAS 12-Month Improvement Outcome (n = 29)** |  |  |
| --- | --- | --- |
|  | **Association with test** | **P** |
| Sex | $X^2=0.0526$ | 0.8185 |
| Race | $X^2=0.7386$ | 0.3901 |
| Ethnicity | $X^2=0.5655$ | 0.4520 |
| Baseline VR-12 score over or under 40 | $X^2=2.3549$ | 0.1249 |
| Tear size | $X^2=1.3773$ | 0.2406 |
| PT visit frequency | $X^2=5.3089$ | 0.3794 |
| Age | F=0.30 | 0.5912 |
| Baseline VAS | F=12.19, 0.6 per preVAS point,<br>$R^2=0.31$ | 0.0017* |
| Baseline ASES Function | F=0.08 | 0.7783 |
| Baseline VR-12 MCS | F=0.28 | 0.5991 |
| PT evaluation wait time | F=1.05 | 0.3140 |
| Number of PT visits | F=0.01 | 0.9067 |
| ASES function improvement | F=139.21 | <0.0001 |
| **ASES Function 12-Month Improvement Outcome (n = 24)** |  |  |
| Sex | $X^2=0.0763$ | 0.7823 |
| Race | $X^2=0.0110$ | 0.9166 |
| Ethnicity | $X^2=0.2757$ | 0.5995 |
| Baseline VR-12 score over or under 40 | $X^2=0.00$ | 1.000 |
| Tear size | $X^2=0.4431$ | 0.5056 |
| PT visit frequency | $X^2=3.2025$ | 0.6688 |
| Age | F=0.02 | 0.8860 |
| Baseline VAS | F=1.26 | 0.2745 |
| Baseline ASES Function | F=9.01, -0.85 per pre-ASES point,<br>$R^2=0.29$ | 0.0066* |
| Baseline VR-12 MCS | F=0.05 | 0.8205 |
| PT evaluation wait time | F=0.46 | 0.5048 |
| Number of PT visits | F=0.02 | 0.8992 |

\**P* value under 0.05
ASES = American Shoulder and Elbow Surgeons, PT = physical therapy, VAS = visual analog scale, VR-12 MCS = Veterans RAND 12-Item Health Survey Mental Component Score

The 12-month ASES function improvement was significantly but inversely associated with baseline ASES functional scores (F = 9.01, *P* = 0.0066). With higher baseline function, less functional improvement is predicted. At the individual level, a lesser 12-month ASES function improvement by 0.85 points can be expected per every 1.0 point increase in baseline ASES score.

### Multivariable Analyses

The multivariable model independent variables included baseline VAS or ASES function, age, VR-12 MCS, tear size, race, ethnicity, sex, evaluation wait time, and total number of PT visits. None of the four models’ results were statistically significant, and none of the independent variables besides baseline VAS and ASES function emerged as a significant predictor while accounting for the others.

## DISCUSSION

The primary objective of this study was to identify the postoperative PT utilization factors that are associated with 12-month results following RCR surgery. Our participants had a mean 12- month VAS improvement of 3.5 and a mean ASES function improvement of 12.1. To our knowledge, there are no previous studies of outcomes of postoperative physical therapy treatment utilization at the clinic of an academic medical center. Stern et al^3^ recommended future research to better understand predictors and variation in outcomes of postoperative RCR rehabilitation, which we have started to build on with this manuscript. We found that with univariate analysis in our sample, 12-month results and improvements were predicted by baseline PRO measures, and not demographic characteristics or PT utilization factors.

An earlier study examined the correlations between baseline outcomes and 1-year postoperative pain and PRO measures in 93 patients undergoing RCR.^29^ We built on this study by including PT utilization data comparing 12-month improvements and attempting a multivariable analysis. Similar to our findings of the baseline scores predicting 12-month VAS scores, these authors found the 12-month VAS correlated with baseline VAS, but not baseline ASES. We also did not find age, sex, or tear size to correlate with 12-month postoperative pain scores.^29^ They found baseline VAS to be correlated with composite 12-month ASES, in contrast to our findings, but our analysis contained only the functional index while theirs included the composite ASES pain measures. In another study of major healthcare utilization outcomes (undergoing repeat RCR and developing capsulitis), no significant associations were found with these outcomes and rehabilitation initiation timing.^12^ Our study examined a similar theme but with inclusion of more granular, individual patient-level PROs.

### Clinical Implications

Although we are not yet ready to make recommendations on PT treatment planning utilizations, we are comfortable applying these findings by using baseline scores to predict 12-month absolute and improvement PRO measures. At the individual level, a higher 12-month VAS score of 0.4 can be predicted by every 1.0 point increase in baseline VAS score. Patients with high preoperative pain levels can be counseled that a higher absolute pain level is to be expected 12 months postoperatively. With higher baseline pain levels, greater pain improvement is also to be expected. A greater 12-month VAS improvement of 0.6 can be expected per every 1.0 point increase in baseline VAS scores. To identify the patients likely to reach an MCID of 2.5, a baseline VAS score of 4.2 or greater would be needed.

In contrast to the relationship with baseline pain levels, we found that a better baseline functional score is predicted to have lower improvement. A lower 12-month ASES functional improvement of 0.85 can be expected per every 1.0 point increase in baseline ASES functional score. This may be a result of the ASES functional score’s ceiling effect, in which those with high preoperative scores may not have the need to improve significantly further to reach their functional goals.

### Multivariable Analysis

Although attempted, the results of our data in multivariable models do not appear to be valid. For the multivariable models, our subjects per variable ratio was 3.2, which may indicate overfitting. During the study’s planning, we did not perform a sample size analysis but decided to screen consecutive cases treated over a pre-specified time period. We did not estimate that so many of our potential participants would be missing 12-month PRO data, and thus we expected a far higher subject per variable ratio.

### Limitations

With this study, we were unable to evaluate all modifiable and non-modifiable risk factors known to affect recovery. There are other clinical comorbid conditions shown to affect RCR outcomes, including low bone mineral density^30^ or diabetes mellitus.^20^ We did not have imaging data at the 12-month follow-up and could not examine rate of re-tear or development of other structural pathology such as glenohumeral osteoarthritis. With data collected via the Epic electronic medical records system, we could only assess participants treated at our institution’s associated therapy clinic. With inclusion of participants treated at an outside clinic, we may have had a larger sample size but would not have been able to ensure that the outside clinics used the same standardized rehabilitation protocol. It is also possible that our participants received PT for other comorbid disorders such as neck pain during the first 6 months following surgery. We also did not monitor home exercise program compliance.

### Future Research

Future research should assess the magnitude and timing of greatest gains in range of motion and strength, as these are important outcomes in addition to pain relief.^31^ Future work of the same design but with a higher sample size may potentially find significant predictors with multivariable analysis.

## Data Availability

All data produced in the present study are available upon reasonable request to the corresponding author.

## Acknowledgements

We would like to thank our enthusiastic participants for volunteering for this study. We also acknowledge David Houghton, Ph.D. for his contributions to planning this project.

## Author Contributions

Author CJP, TE, and JS designed research. JS and BP conducted research and collected data. CJP and JS developed statistical analysis, and CJP led data analysis. CJP, TE, BP, JCH, and JS participated in manuscript writing and revision. JS had primary responsibility for the final manuscript. All authors have read and approved the final manuscript.

## Data Sharing Statement

The datasets generated during and/or analyzed during the current study are available from the corresponding author on reasonable request.

## Funding

This study received no additional funding.

## Conflicts of Interest

Chris J. Pierson-no conflicts of interest.

Brady Moore-no conflicts of interest.

Tristan Elias-no conflicts of interest.

Joshua C. Harris-no conflicts of interest.

Jeremy Somerson is a paid consultant for Exactech, a stockholder in Johnson and Johnson, Lilly, Intuitive Surgical, United Healthcare, and provides educational support for Medinc of Texas.

## List of abbreviations

AAOS: American Academy of Orthopaedic Surgeons
ASES: American Shoulder and Elbow Surgeons
ICD-9: International Classification of Diseases, Ninth Revision
PT: Physical Therapy
PRO: Patient-Reported Outcome Measure
RCR: Rotator Cuff Repair
VAS: Visual Analog Scale
VR-12 MCS: Veterans RAND 12-item Health Survey Mental Component Summary score

## Notes

### Funding Statement

This study did not receive any funding.

### Author Declarations

This study received Institutional Review Board approval (IRB# 20-0306) at the University of Texas Medical Branch.

## REFERENCES

1. Zhang AL, Montgomery SR, Ngo SS, Hame SL, Wang JC, Gamradt SC. Analysis of rotator cuff repair trends in a large private insurance population. Arthroscopy. Apr 2013;29(4):623–9. doi:10.1016/j.arthro.2012.11.004

2. Claes A, Mertens MG, Verborgt O, Baert I, Struyf F. Factors associated with better treatment outcome of physical therapy interventions after shoulder arthroplasty: A systematic review. Clin Rehabil. Oct 2022;36(10):1369–1399. doi:10.1177/02692155221106627

3. Stern BZ, Zubizarreta N, Anthony SG, Gladstone JN, Poeran J. Variation in utilization of physical therapist and occupational therapist services after rotator cuff repair: a population-based study. Phys Ther. Apr 2 2024;104(4)doi:10.1093/ptj/pzae015

4. Mall NA, Tanaka MJ, Choi LS, Paletta GA, Jr. Factors affecting rotator cuff healing. J Bone Joint Surg Am. May 7 2014;96(9):778–88. doi:10.2106/jbjs.M.00583

5. Mather RC, 3rd, Koenig L, Kocher MS, et al. Societal and economic impact of anterior cruciate ligament tears. J Bone Joint Surg Am. Oct 2 2013;95(19):1751–9. doi:10.2106/jbjs.L.01705

6. Ross D, Maerz T, Lynch J, Norris S, Baker K, Anderson K. Rehabilitation following arthroscopic rotator cuff repair: a review of current literature. J Am Acad Orthop Surg. Jan 2014;22(1):1–9. doi:10.5435/jaaos-22-01-1

7. Arce G, Bak K, Bain G, et al. Management of disorders of the rotator cuff: proceedings of the ISAKOS upper extremity committee consensus meeting. Arthroscopy. Nov 2013;29(11):1840–50. doi:10.1016/j.arthro.2013.07.265

8. Ghodadra NS, Provencher MT, Verma NN, Wilk KE, Romeo AA. Open, mini-open, and all-arthroscopic rotator cuff repair surgery: indications and implications for rehabilitation. J Orthop Sports Phys Ther. Feb 2009;39(2):81–9. doi:10.2519/jospt.2009.2918

9. Papalia R, Franceschi F, Zampogna B, D’Adamio S, Maffulli N, Denaro V. Augmentation techniques for rotator cuff repair. Br Med Bull. 2013;105:107–38. doi:10.1093/bmb/lds029

10. Arshi A, Kabir N, Cohen JR, et al. Utilization and costs of postoperative physical therapy after rotator cuff repair: a comparison of privately insured and Medicare patients. Arthroscopy. Dec 2015;31(12):2392–9.e1. doi:10.1016/j.arthro.2015.06.018

11. Baumann A, Indermuhle T, Curtis D, Perez J, Leland JM, 3rd. Factors affecting postoperative rehabilitation therapy utilization after arthroscopic rotator cuff repair: an epidemiological analysis. Cureus. Mar 2023;15(3):e36740. doi:10.7759/cureus.36740

12. Stern BZ, Zubizarreta N, Anthony SG, Poeran J, Gladstone JN. Association between timing of initiating supervised physical rehabilitation after rotator cuff repair and incidence of repeat repair and capsulitis: a population-based analysis. J Shoulder Elbow Surg. Aug 2024;33(8):1747–1754. doi:10.1016/j.jse.2024.01.017

13. von Elm E, Altman DG, Egger M, Pocock SJ, Gøtzsche PC, Vandenbroucke JP. Strengthening the Reporting of Observational Studies in Epidemiology (STROBE) statement: guidelines for reporting observational studies. BMJ. Oct 20 2007;335(7624):806–8. doi:10.1136/bmj.39335.541782.AD

14. Thigpen CA, Shaffer MA, Gaunt BW, Leggin BG, Williams GR, Wilcox RB, 3rd. The American Society of Shoulder and Elbow Therapists’ consensus statement on rehabilitation following arthroscopic rotator cuff repair. J Shoulder Elbow Surg. Apr 2016;25(4):521–35. doi:10.1016/j.jse.2015.12.018

15. American Academy of Orthopaedic Surgeons. Management of rotator cuff injuries: evidenced-based clinical practice guideline. Accessed October 21, 2024. https://www.aaos.org/rccpg

16. Richards RR, An KN, Bigliani LU, et al. A standardized method for the assessment of shoulder function. J Shoulder Elbow Surg. Nov 1994;3(6):347–52. doi:10.1016/s1058-2746(09)80019-0

17. Michener LA, McClure PW, Sennett BJ. American Shoulder and Elbow Surgeons Standardized Shoulder Assessment Form, patient self-report section: reliability, validity, and responsiveness. J Shoulder Elbow Surg. Nov-Dec 2002;11(6):587–94. doi:10.1067/mse.2002.127096

18. Boileau P, Brassart N, Watkinson DJ, Carles M, Hatzidakis AM, Krishnan SG. Arthroscopic repair of full-thickness tears of the supraspinatus: does the tendon really heal? J Bone Joint Surg Am. Jun 2005;87(6):1229–40. doi:10.2106/jbjs.D.02035

19. Cho NS, Rhee YG. The factors affecting the clinical outcome and integrity of arthroscopically repaired rotator cuff tears of the shoulder. Clin Orthop Surg. Jun 2009;1(2):96–104. doi:10.4055/cios.2009.1.2.96

20. Chung SW, Park JS, Kim SH, Shin SH, Oh JH. Quality of life after arthroscopic rotator cuff repair: evaluation using SF-36 and an analysis of affecting clinical factors. Am J Sports Med. Mar 2012;40(3):631–9. doi:10.1177/0363546511430309

21. Gulotta LV, Nho SJ, Dodson CC, Adler RS, Altchek DW, MacGillivray JD. Prospective evaluation of arthroscopic rotator cuff repairs at 5 years: part II--prognostic factors for clinical and radiographic outcomes. J Shoulder Elbow Surg. Sep 2011;20(6):941–6. doi:10.1016/j.jse.2011.03.028

22. Manaka T, Ito Y, Matsumoto I, Takaoka K, Nakamura H. Functional recovery period after arthroscopic rotator cuff repair: is it predictable before surgery? Clin Orthop Relat Res. Jun 2011;469(6):1660–6. doi:10.1007/s11999-010-1689-6

23. Rizvi SMT, Bishop M, Lam PH, Murrell GAC. Factors predicting frequency and severity of postoperative pain after arthroscopic rotator cuff repair surgery. Am J Sports Med. Jan 2021;49(1):146–153. doi:10.1177/0363546520971749

24. Cuff DJ, O’Brien KC, Pupello DR, Santoni BG. Evaluation of factors affecting acute postoperative pain levels after arthroscopic rotator cuff repair. Arthroscopy. Jul 2016;32(7):1231–6. doi:10.1016/j.arthro.2015.12.021

25. Moore BP, Forrister DZ, Somerson JS. A threshold of lower preoperative mental health is associated with decreased achievement of comfort and capability benchmarks following rotator cuff repair: a retrospective cohort study. J Shoulder Elbow Surg. Aug 2024;33(8):e403–e414. doi:10.1016/j.jse.2023.12.011

26. Martin JR, Castaneda P, Kisana H, McKee MD, Amini MH. Preoperative patient-reported outcomes predict postoperative clinical outcomes following rotator cuff repair. Arthroscopy. May 2024;40(5):1445–1452. doi:10.1016/j.arthro.2023.10.008

27. Yeo DY, Walton JR, Lam P, Murrell GA. The relationship between intraoperative tear dimensions and postoperative pain in 1624 consecutive arthroscopic rotator cuff repairs. Am J Sports Med. Mar 2017;45(4):788–793. doi:10.1177/0363546516675168

28. Dworkin RH, Turk DC, Wyrwich KW, et al. Interpreting the clinical importance of treatment outcomes in chronic pain clinical trials: IMMPACT recommendations. J Pain. Feb 2008;9(2):105–21. doi:10.1016/j.jpain.2007.09.005

29. Ravindra A, Barlow JD, Jones GL, Bishop JY. A prospective evaluation of predictors of pain after arthroscopic rotator cuff repair: psychosocial factors have a stronger association than structural factors. J Shoulder Elbow Surg. Oct 2018;27(10):1824–1829. doi:10.1016/j.jse.2018.06.019

30. Chung SW, Oh JH, Gong HS, Kim JY, Kim SH. Factors affecting rotator cuff healing after arthroscopic repair: osteoporosis as one of the independent risk factors. Am J Sports Med. Oct 2011;39(10):2099–107. doi:10.1177/0363546511415659

31. Karpinski K, Plachel F, Gerhardt C, et al. Different expectations of patients and surgeons with regard to rotator cuff repair. J Shoulder Elbow Surg. May 2022;31(5):1096–1105. doi:10.1016/j.jse.2021.12.043

